# T-cell activation and fibroblastic BMP4-Gremlin dysregulation indicate disease severity in acute myocarditis

**DOI:** 10.64898/2026.04.10.26350598

**Authors:** Anna Joachimbauer, Christian Perez-Shibayama, Emily Payne, Isabella Hanka, Roman Stadler, Iliana Papadopoulou, Hans Rickli, Micha T. Mäder, Oliver Borst, Monika Zdanyte, Leslie Cooper, Lukas Flatz, Christian M. Matter, Verena C. Wilzeck, Robert Manka, Ardan M. Saguner, Frank Ruschitzka, Dörthe Schmidt, Burkhard Ludewig, Cristina Gil-Cruz

## Abstract

**Background and Aims:** Acute myocarditis (AM) is a T cell-mediated myocardial disease with clinical manifestations ranging from mild chest pain to cardiogenic shock. Reliable biomarkers to stratify patients and guide therapy are currently lacking. In particular, the extent of the dysregulation of inflammatory pathways, and the impact on myocardial dysfunction, remain elusive.

**Methods:** Serum analyses were performed in prospectively recruited AM patients (n = 103) from two independent cohorts. Multimodal data integration combining profiling of cytokine and chemokine dysregulation with clinical biomarkers was used to define clinical phenotypes with distinct inflammatory signatures. Machine-learning and regression models were applied to determine biomarkers that indicate clinical severity.

**Results:** Immuno-proteomic profiling revealed conserved inflammatory patterns across AM cohorts, dominated by T cell-related cytokines and chemokines. In addition, AM patients showed dysregulation of fibroblast-derived cytokines, including hepatocyte growth factor (HGF), bone morphogenic protein 4 (BMP4) and the BMP4 inhibitors Gremlin-1 (GREM1) and Gremlin-2 (GREM2). Data integration and unsupervised clustering revealed two immuno-clinical phenotypes, linking T cell activation and fibroblast dysregulation to disease severity. Machine learning-based analysis identified CXCL10, GREM2 and LVEF as critical parameters for stratifying disease severity.

**Conclusions:** These findings highlight a systemic T cell activation signature as diagnostic hallmark of AM. In addition, dysregulation of fibroblast-derived tissue cytokines serves as an indicator for distinct immuno-clinical phenotypes in myocardial inflammatory disease. Thus, the clinically relevant link between T cell-driven immune activation, myocardial inflammation and fibroblast-driven remodelling provides a versatile set of parameters to identify severe manifestations of AM.

**Graphical Abstract:** **Key Question:** Are serological immune signatures linked to clinical severity in acute myocarditis and do they enable patient stratification?

**Key Findings:** T cell- and fibroblast associated proteomic signatures indicate disease severity in acute myocarditis. Novel immuno-clinical phenotypes stratify patients according to distinct immune responses and clinical manifestations. CXCL10, GREM2 and LVEF are the most important parameters to identify immuno-clinical phenotypes.

**Take Home Message:** CXCL10, GREM2 and LVEF emerge as key determinants for a severe immuno-clinical phenotype in acute myocarditis, highlighting the role of T cell-fibroblast interaction in the disease process and linking T cell activation, fibroblastic tissue remodelling and impaired cardiac function.

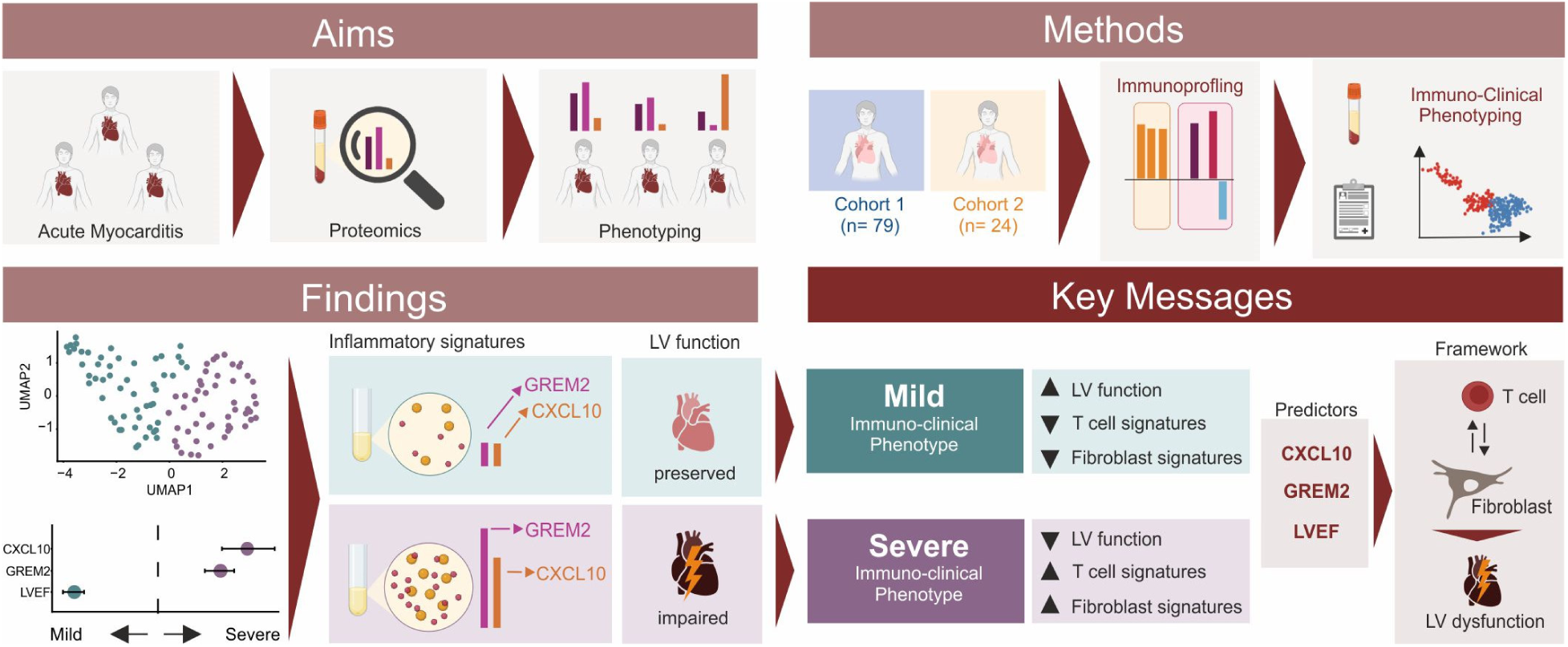

## Introduction

Acute myocarditis (AM) is an inflammatory disease of the myocardium that presents as a heterogeneous clinical entity, with symptoms ranging from isolated chest pain to cardiogenic shock ^1–3^. However, the common immunopathological mechanisms underlying the broad spectrum of disease manifestations remain poorly understood. As the current risk stratification in AM relies predominantly on unspecific clinical parameters ^2,3^, there is an apparent need for biomarkers that reflect the inflammatory nature of the disease. Furthermore, an improved mechanistic understanding of AM pathogenesis and the immunological processes driving clinical manifestations could facilitate patient stratification and guide therapeutic strategies.

While viral infections are the most common trigger of myocarditis ^4^, the frequent absence of viral genomes in endomyocardial biopsies points to T cell-driven immune processes as the primary mechanism driving disease ^2,5,6^. Indeed, the identification of heart-specific T cells ^7–10^ and the activation of T cells following immune checkpoint inhibitor (ICI) therapy ^11–13^ support the conclusion that T cells are the cause for progressive inflammation in AM. Autoimmune T cells with cardiopathogenic potential are activated through viral infection-associated bystander effects, unleashed during ICI therapy or induced during myocardial damage ^2^. Such potentially cardiotoxic T cells recognize epitopes derived from cardiac antigens including myosin heavy-chain 6 (MYH6) ^8–10,13,14^, Troponin ^15^ or beta-1 adrenergic receptor (ADRB1) ^16,17^. Importantly, the presence of heart-specific T cells in the circulation of healthy individuals highlights the delicate balance of stimulatory and counter-regulatory mechanisms controlling cardiopathogenic T cell activity ^8,14^.

The accumulation of activated T cells in the myocardium drives profound alterations of the cardiac microenvironment and thereby contributes to myocardial damage ^10,18,19^. T cell infiltration induces reprogramming of cardiac fibroblasts toward an inflammatory phenotype, characterized by increased production of proinflammatory cytokines and chemokines that perpetuate pathological processes and loss of myocardial function ^20–22^. Inflammation-driven fibroblast activation is accompanied by enhanced tissue-remodelling activity including increased extracellular matrix (ECM) production and lymphangiogenesis ^22–24^. Intercellular crosstalk between activated T cells and fibroblasts is dominated by T-helper 1 and 17 pathways ^9,25–27^ and is modulated primarily by myeloid cell-derived cytokines, such as IL-6 and IL-1 ^28,29^. In addition, autocrine regulation of cardiac fibroblasts via bone morphogenetic protein 4 (BMP4) and its inhibitors Gremlin-1 (GREM1) and Gremlin-2 (GREM2) contributes to tissue homeostasis and cardiac remodelling ^22^. The critical role of BMP4 for balancing homeostatic and inflammatory processes in the myocardium is reflected by altered BMP4 concentration in the serum of AM patients ^22^. It is thus important to assess the extent of altered myocardial fibroblast-T cell interactions in AM patients and to identify a distinct set of immuno-clinical parameters to improve patient stratification.

Here, we provide serological immuno-proteomic signatures that capture immune activation and inflammation-induced tissue remodelling processes in AM patients. The integration of comprehensive immunologic profiling with multimodal clinical data facilitated the distinction of immuno-clinical profiles that guide the assessment of clinical severity in AM. Increased expression of the T cell chemokine CXCL10 and the modulator of fibroblast activity GREM2 in conjunction with reduced left ventricular ejection fraction (LVEF) emerge as key determinants for a severe immuno-clinical phenotype. These data underpin the critical role of T cell-fibroblast interaction in acute myocardial inflammation and link T cell activation to fibroblastic tissue remodelling and impaired cardiac function.

## Methods

### Study Cohorts and sample collection

The main study cohort (Cohort 1) was derived from the ImmpathCarditis study (ethical approval: BASEC 2021-01917), a prospective longitudinal study recruiting patients with AM between May 2022 and November 2025 at the University Hospital Zürich and the Cantonal Hospital St. Gallen. Inclusion criteria required at least one clinical symptom and one diagnostic criterion ^3^, confirmation of AM by cardiac magnetic resonance (CMR) according to the updated Lake Louise Criteria ^30^ or by endomyocardial biopsy (EMB) and age 16-85 years. Exclusion criteria included the history of autoimmune or chronic infectious diseases (e.g., SLE, Lyme disease, HIV, HBV or HCV) and significant coronary artery disease (>50% stenosis on invasive or computed tomography coronary angiography). An external, independent cohort of AM patients was included (Cohort 2). Patients were prospectively recruited at the University Hospital Tübingen between January 2023 and August 2025 (ethical approval: 236/2018BO2). Diagnosis of AM was confirmed by EMB and/or by CMR in accordance with current myocarditis guidelines ^3^. For cross-disease validation, patients with arrhythmogenic cardiomyopathy (ACM) were enrolled at the University Hospital Zürich through the multicentre ACM registry and biobank between March 2022 and July 2023 (ethical approval: 2014-0443) ACM was diagnosed according to the 2010 revised Task Force Criteria. ^31^ Healthy controls were recruited through the IMIT study (ethical approval: 2016-00998). Participants had no prior history of cardiomyopathies, myocarditis, chronic inflammatory or autoimmune diseases and no infectious symptoms within four weeks prior to sampling. Controls were age- and sex-matched to the main study cohort.

All studies were approved by local ethics committees and conducted in accordance with the declaration of Helsinki. Clinical data were extracted from the institutional patient information systems across all cohorts. Cardiac functional parameters were derived from the first echocardiographic assessment after admission in the AM cohorts or from the most recent echocardiographic assessment in the ACM cohort (mean interval of 5.2 months relative to blood sampling). Blood samples from the AM cohorts were collected at study enrolment (Cohort 1: median 3 days after admission, Cohort 2: median 2 days after admission) and samples from the ACM cohort were obtained during routine follow-up. All blood samples were collected in serum tubes, centrifuged at 1,500 rpm for 10 min, aliquoted into cryovials and stored at −80°C until analysis.

### Routine blood parameters

In both AM cohorts, routine blood parameters, including C-reactive protein (CRP), Troponin I and N-terminal pro-B-type natriuretic peptide (NT-proBNP) are reported as peak values recorded during hospitalisation. In the healthy control and ACM cohorts routine blood parameters were measured from study-specific blood samples.

### Luminex Multiplex Assay

Undiluted serum samples were measured to quantify the concentration of cytokines, chemokines and growth factors using the Cytokine 30-Plex Human Panel (Invitrogen). Samples were measured upon first thawing. Assay plate preparation, including the dilution of standard curves, was conducted according to the assay instructions of the manufacturer. Plates were subsequently read on Luminex instruments (FLEXMAP 3D xPONENT 4.2 System). Mean fluorescence intensity values of each analyte were recorded and concentrations were calculated based on standard curves generated using a five-parameter logistic weighted regression model implemented in the manufacturer’s software. Values below the lower limit of detection or above the upper limit of detection were assigned to the respective detection limit values of the corresponding biomarker. A complete list of biomarkers and their detection limits is provided in the Supplementary data, Table 1. Serum samples were assayed in duplicates and the mean of the duplicate measurements was used for subsequent analyses. For quality control and further inter-assay comparison, a positive control was included in each assay run. Quantified serum protein concentrations were statistically analysed using R (Version 4.4.3).

### BMP4, GREM1 and GREM2 ELISA

BMP4 serum concentrations were measured using a commercial BMP4 ELISA Kit (Abcam) and samples were diluted 1:4 in Sample Diluent NS. The detection limit of the assay is 2.5 pg/ml. GREM1 serum concentrations were measured with the Gremlin DuoSet ELISA (R&D Systems). GREM2 serum concentrations were detected using the Mouse PRDC/GREM2 DuoSet ELISA (R&D Systems). Human and mouse GREM2 share approximately 94% amino acid homology allowing for cross-species detection ^32,33^. All assays were performed according to the manufacturer’s instructions. Measurements were done in duplicates and results are reported as mean value.

### Statistical Analysis

Continuous data are presented as individual data points with mean, standard deviation (SD) for normally distributed data or median and interquartile range for non-normally distributed data. Categorical data are reported as counts and percentages. Associations are expressed as odds ratios (OR) with a 95% confidence interval (CI). Missing data were imputed using multivariate imputation by chained equations (MICE). The multivariate imputation was performed with 5 iterations of the imputation process and created 5 multiple imputations data sets. Imputation plausibility was assessed by comparing density distributions of observed and imputed values ^34^. Normality was evaluated using the Shapiro-Wilk test, indicating non-parametric distributions in our dataset. The non-parametric Mann-Whitney U test was used for comparison between two groups and the Kruskal-Wallis test with Dunn’s post hoc correction for multiple groups. Log2 fold changes were derived from non-parametric comparisons. Correlations were assessed using the non-parametric Spearman correlation. Hierarchical clustering in heatmap visualization was performed using Ward’s method. Dimensionality reduction of combined clinical and immunological data was conducted using Uniform Manifold Approximation and Projection (UMAP). Prior to dimensionality reduction and unsupervised clustering, parameters were preselected to reduce noise and enhance cluster stability including only parameters exhibiting a log2 fold change (FC) >1 in at least 50% of AM patients. Immuno-clinical phenotypes were identified by unsupervised spiral clustering to the dimension-reduced data. To identify variables predictive of immuno-clinical phenotypes, a random forest model was trained using the previously identified immuno-clinical phenotypes as the outcome variable. Model training and evaluation were performed using the caret R package. Ten-fold cross-validation was applied to assess model performance, by repeatedly splitting the dataset into training (90%) and testing (10%) subsets. The random forest model was trained using 500 trees and tree depth was constrained to a maximum of four terminal nodes to reduce model overfitting. Model accuracy was derived from cross-validated results. Variable importance was assessed using permutation-based importance scores, which quantify the contribution of each variable to model performance. To quantify the effect size and the direction of associations between individual variables and the immuno-clinical phenotypes, we performed ridge regression using the gmlnet R package. The optimal penalty parameter (lambda) was determined via five-fold cross-validation. Confidence intervals and p-values were estimated using 100 bootstrap resampling recapitulations. Model coefficients were expressed as odds ratios (ORs) with corresponding 95% confidence intervals. All statistical analyses were conducted in R version 4.4.3 (R Foundation for Statistical Computing, Vienna, Austria). Statistical significance was defined as p < 0.05. To control for multiple testing, p values were adjusted using the Benjamini-Hochberg method to reduce the false discovery rate (FDR).

### Code availability

The code used for the analysis of this study is available on GitHub: https://ludewig-lab.github.io/Serology-Analysis/

## Results

### Clinical parameters associated with disease severity in acute myocarditis

A total of 103 AM patients were included in this study, recruited from two independent cohorts: Cohort 1 (n = 79) from the University Hospital Zürich and Cantonal Hospital St.Gallen and Cohort 2 (n = 24) from the University Hospital of Tübingen (Figure 1A). Baseline characteristics are summarized in Table 1. In both cohorts, the majority of patients were male (84% in Cohort 1 and 67% in Cohort 2) and the median age was 29 years in Cohort 1 and 48 years in Cohort 2. Cohort 2 included a significantly higher proportion of patients that required mechanical circulatory support (33% vs. 3.8% in Cohort 1, p<0.001) and immunosuppressive therapy (64% vs. 13% in Cohort 1, p<0.001). LVEF was significantly lower in patients of Cohort 2 (median LVEF 31% vs. 53% in Cohort 1, p<0.001; Table 1; Supplementary data, Figure 1A), indicating that Cohort 2 comprised patients with more severe clinical manifestations. Routine blood parameters demonstrated significantly higher NT-proBNP levels in patients of Cohort 2, whereas Troponin I and CRP levels did not differ between cohorts (Supplementary data, Figure 1B-D). Data from both cohorts were pooled for subsequent analyses and sex- and age-matched healthy individuals were used as controls (Figure 1A; Supplementary data, Table 2). AM patients exhibited a broad spectrum of clinical severity, as evidenced by a wide range of LVEF, spanning from preserved to severe impaired cardiac function (Figure 1B). As expected, peak serum levels of Troponin I, CRP and NT-proBNP values were significantly elevated in AM patients when compared to healthy controls (Figure 1C-E). Correlation analyses between cardiac function and routine blood parameters showed that peak Troponin I concentration in serum did not correlate with LVEF (Figure 1F) or NT-proBNP levels (Supplementary data, Figure 1E). While CRP concentration was weakly associated with disease severity (Figure 1G; Supplementary data, Figure 1F), peak NT-proBNP strongly correlated with progressive loss of left ventricular function (Figure 1H), underscoring that LVEF and NT-proBNP represent robust clinical parameters indicating disease severity in AM.

**Figure 1.**
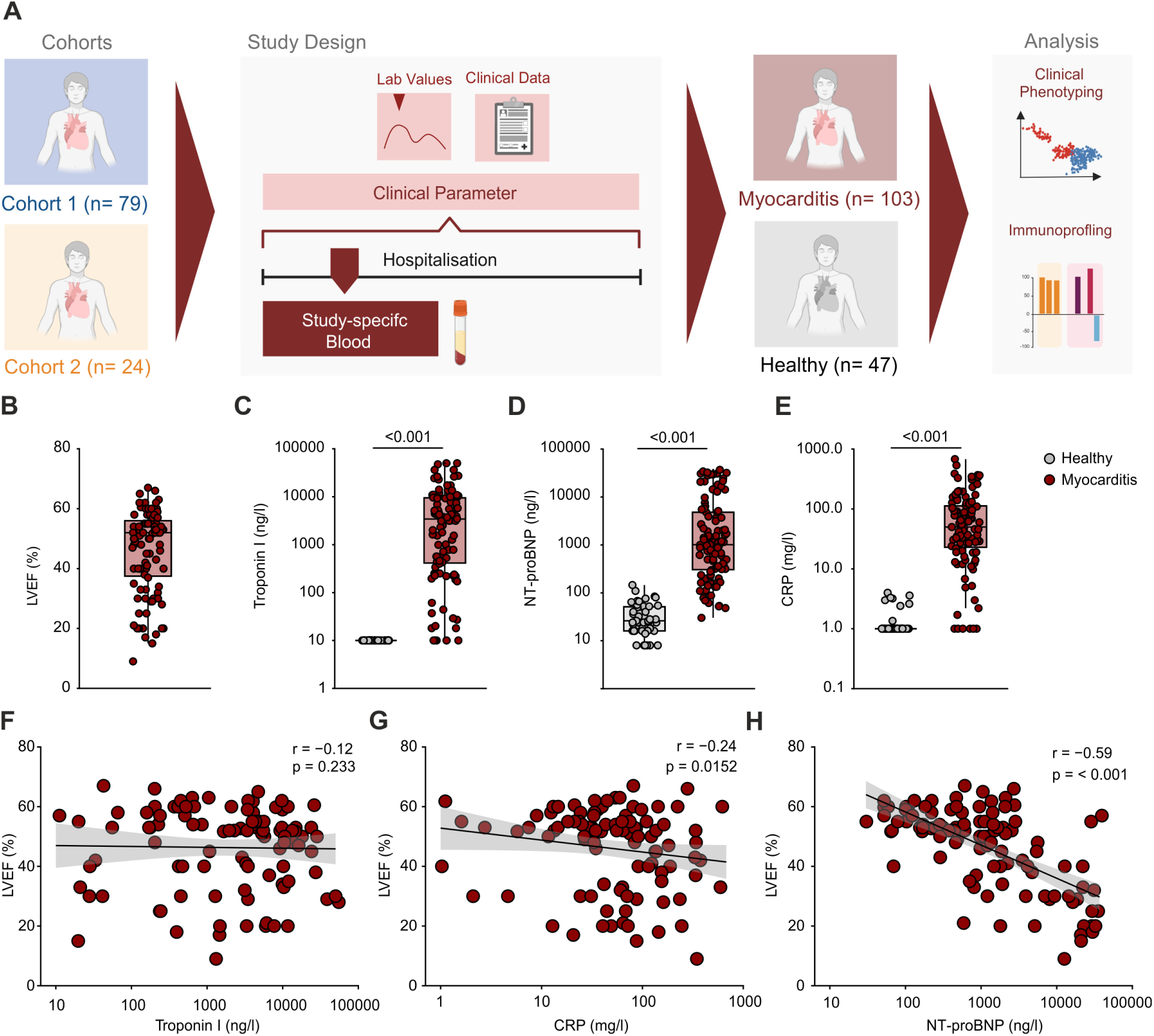
Association of standard clinical parameters with disease severity in acute myocarditis. *(A)* Study design and analytical workflow. (*B)* LVEF in AM patients (n= 103). *(C-E)* Peak values of routine blood parameters during hospitalization: *(C)* Troponin I, *(D)* CRP, *(E)* NT-proBNP. *(F-H)* Correlations between LVEF and routine blood parameters: *(F)* Troponin I, *(G)* CRP, *(H)* NT-proBNP. *(B-H)* Dots represent individual patients, *(B-E)* box and whiskers indicate minimum to maximum values, median, and interquartile range, *(F-H)* linear regression lines with 95% confidence interval (grey shading). Statistical analyses were performed using the Mann-Whitney U test for *(C-E)* and non-parametric Spearman correlation for *(F-H)*. P-values were corrected for multiple testing using the Benjamini-Hochberg method. AM, acute myocarditis, LVEF, left ventricular ejection fraction, CRP, C-reactive protein, NT-proBNP, N-terminal pro-B-type natriuretic peptide.

**Table 1:**
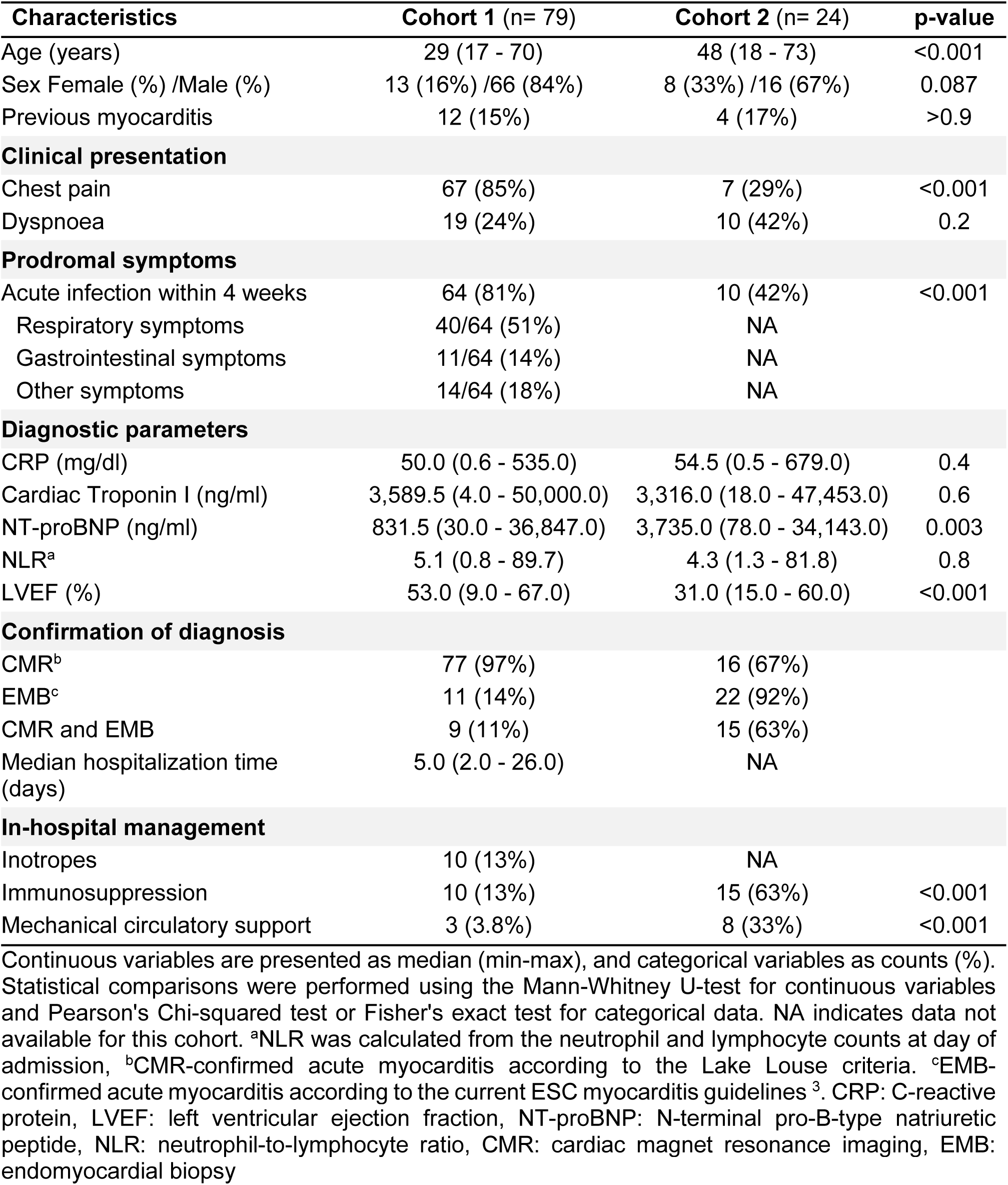
Baseline clinical characteristics of two independent acute myocarditis cohorts.

| Characteristics | Cohort 1 (n= 79) | Cohort 2 (n= 24) | p-value |
| --- | --- | --- | --- |
| Age (years) | 29 (17 - 70) | 48 (18 - 73) | <0.001 |
| Sex Female (%) /Male (%) | 13 (16%) /66 (84%) | 8 (33%) /16 (67%) | 0.087 |
| Previous myocarditis | 12 (15%) | 4 (17%) | >0.9 |
| **Clinical presentation** |  |  |  |
| Chest pain | 67 (85%) | 7 (29%) | <0.001 |
| Dyspnoea | 19 (24%) | 10 (42%) | 0.2 |
| **Prodromal symptoms** |  |  |  |
| Acute infection within 4 weeks | 64 (81%) | 10 (42%) | <0.001 |
| Respiratory symptoms | 40/64 (51%) | NA |  |
| Gastrointestinal symptoms | 11/64 (14%) | NA |  |
| Other symptoms | 14/64 (18%) | NA |  |
| **Diagnostic parameters** |  |  |  |
| CRP (mg/dl) | 50.0 (0.6 - 535.0) | 54.5 (0.5 - 679.0) | 0.4 |
| Cardiac Troponin I (ng/ml) | 3,589.5 (4.0 - 50,000.0) | 3,316.0 (18.0 - 47,453.0) | 0.6 |
| NT-proBNP (ng/ml) | 831.5 (30.0 - 36,847.0) | 3,735.0 (78.0 - 34,143.0) | 0.003 |
| NLR<sup>a</sup> | 5.1 (0.8 - 89.7) | 4.3 (1.3 - 81.8) | 0.8 |
| LVEF (%) | 53.0 (9.0 - 67.0) | 31.0 (15.0 - 60.0) | <0.001 |
| **Confirmation of diagnosis** |  |  |  |
| CMR<sup>b</sup> | 77 (97%) | 16 (67%) |  |
| EMB<sup>c</sup> | 11 (14%) | 22 (92%) |  |
| CMR and EMB | 9 (11%) | 15 (63%) |  |
| Median hospitalization time (days) | 5.0 (2.0 - 26.0) | NA |  |
| **In-hospital management** |  |  |  |
| Inotropes | 10 (13%) | NA |  |
| Immunosuppression | 10 (13%) | 15 (63%) | <0.001 |
| Mechanical circulatory support | 3 (3.8%) | 8 (33%) | <0.001 |

Continuous variables are presented as median (min-max), and categorical variables as counts (%). Statistical comparisons were performed using the Mann-Whitney U-test for continuous variables and Pearson's Chi-squared test or Fisher's exact test for categorical data. NA indicates data not available for this cohort. <sup>a</sup>NLR was calculated from the neutrophil and lymphocyte counts at day of admission, <sup>b</sup>CMR-confirmed acute myocarditis according to the Lake Louise criteria. <sup>c</sup>EMB-confirmed acute myocarditis according to the current ESC myocarditis guidelines <sup>3</sup>. CRP: C-reactive protein, LVEF: left ventricular ejection fraction, NT-proBNP: N-terminal pro-B-type natriuretic peptide, NLR: neutrophil-to-lymphocyte ratio, CMR: cardiac magnet resonance imaging, EMB: endomyocardial biopsy

### T cell activation signatures define the immune response of acute myocarditis

As the inflammatory marker CRP showed only a weak association with disease severity (Figure 1G), we used targeted immuno-proteomic serum analyses to identify dominant immune activation signatures associated with AM. Patients from both AM cohorts exhibited a broad upregulation of inflammatory cytokines, chemokines and fibroblast-derived tissue cytokines when compared with healthy controls (Supplementary data, Figure 2A). Increased clinical severity in AM patients from Cohort 2 appeared to be associated with enhanced production of inflammatory cytokines and chemokines (Figure 2A), as reflected by the broad spectrum of significantly upregulated inflammatory molecules (Supplementary data, Figure 2B-C). Comparative analysis of cytokine and chemokine expression between both AM cohorts identified a conserved set of eight inflammatory molecules significantly elevated in all AM patients with a log2 FC >1.5: hepatocyte growth factor (HGF), interleukin-2 receptor (IL-2R), CXCL9, CXCL10, CXCL8, CCL3 and CCL4 (Figure 2B, Supplementary data, Figure 2D-K) Unsupervised hierarchical clustering revealed a dominant T cell-associated inflammatory signature across all AM patients, characterized by elevated T cell attracting chemokines CXCL9 and CXCL10 and the increased production of the T cell activation marker IL-2R (Figure 2C). The dominant T cell signature was accompanied by elevated levels of the fibroblast-derived tissue cytokine HGF (Figure 2C). Interestingly, AM patients showed increased serum levels of additional fibroblast-derived factors such as epithelial growth factor (EGF), fibroblast growth factor-2 (FGF2) and vascular endothelial growth factor-A (VEGF-A) when compared with healthy controls (Supplementary data, Figure 2L-N). In addition to the dominant T cell-associated signature, a subset of patients exhibited elevated levels of inflammatory mediators that are commonly produced in the inflamed myocardium by activated myeloid cells ^28^, i.e., CXCL8, CCL3, CCL4 and IL-6 (Figure 2C; Supplementary data, Figure 2H-K). Correlation analysis revealed the highly significant association of NT-proBNP serum concentration with the T cell-derived markers IL-2R, CXCL9, and CXCL10 and the fibroblast-derived cytokine HGF (Figure 2D-F, Supplementary Figure 2O), while the association of NT-proBNP with myeloid-associated inflammatory mediators was less consistent and not uniformly observed across patients (Supplementary data, Figure 2P-S). Taken together, the dominant inflammatory signature supports the notion that AM is a T cell-driven disease and that T cell-fibroblast crosstalk is linked to disease severity in AM patients.

**Figure 2.**
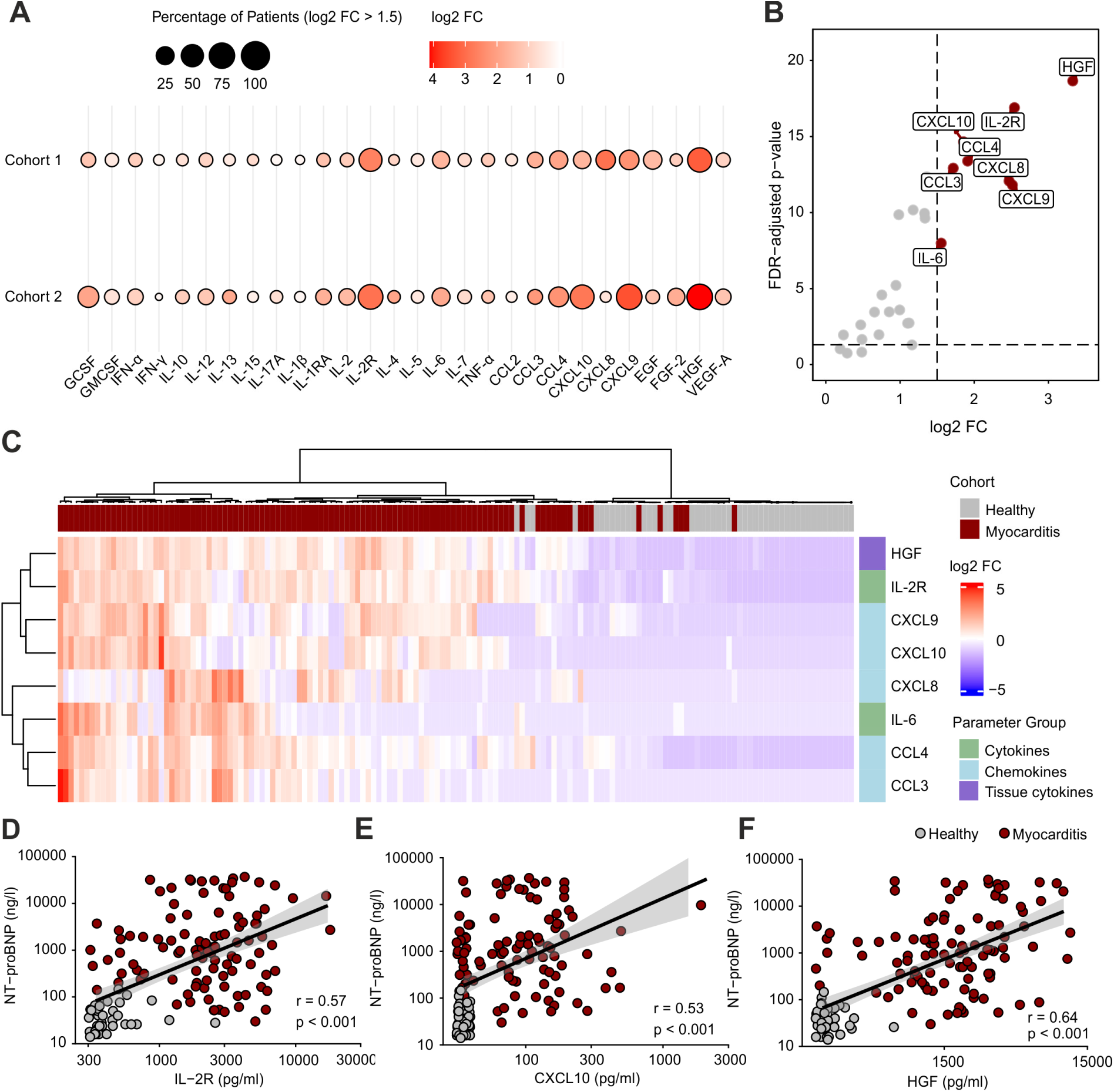
Conserved inflammatory signatures in AM patients. *(A)* Bubble plot showing expression pattern of 28 cytokines, chemokines and tissue-derived cytokines in sera from two independent AM cohorts (Cohort 1 n = 79, Cohort 2 n = 24) relative to healthy controls. *(B)* Volcano plot with differentially expressed proteins in AM patients (n = 103) compared to healthy controls. *(C)* Heatmap of inflammatory serum molecules in AM patients and healthy controls. *(D-F)* Correlation analyses of NT-proBNP and inflammatory molecules: *(D)* IL-2R, *(E)* CXCL10, *(F)* HGF. *(A)* Bubble size represents the abundance of AM patients expressing inflammatory molecules >1.5 log2 FC and bubble colour indicates magnitude of log2 FC. *(B)* Red dots indicate inflammatory molecules expressed >1.5 log2 FC and p <0.05 in AM patients. *(C)* High protein expression is shown in red and low expression in blue, indicated as log2 FC. *(D-F)* Dots represent individual patients, linear regression lines with 95% confidence interval (grey shading). Statistical analyses were performed using the Mann-Whitney U-test for *(A-B),* Ward’s unsupervised hierarchical clustering method for (C), and non-parametric Spearman correlation for *(D-F)*. P-values were corrected for multiple testing using the Benjamini-Hochberg method. AM, acute myocarditis, FC, fold change, G-CSF, granulocyte colony-stimulating factor, GM-CSF, granulocyte-macrophage colony-stimulating factor, IFN-α, interferon alpha, IFN-γ, interferon gamma, IL-1β, interleukin-1 beta, IL-1RA, interleukin-1 receptor antagonist, IL-2R, interleukin-2 receptor, TNF-α, tumor necrosis factor alpha, EGF, epidermal growth factor, HGF, hepatocyte-growth factor, VEGF-A, vascular endothelial growth factor A, FDR, false discovery rate, NT-proBNP, N-terminal pro-B-type natriuretic peptide

### BMP4/GREM2 dysregulation indicates disease severity in acute myocarditis

The pronounced elevation of fibroblast-derived factors in the serum of AM patients and the association of HGF with disease severity, led us to further extend the panel of fibroblast markers that are mechanistically linked to myocardial inflammation in AM ^22^. To this end, we quantified serum concentration of BMP4 and its inhibitors GREM1 and GREM2 in both AM cohorts, in comparison to healthy controls (Figure 3A-C, Supplementary data, Figure 3A-C). Consistent with previously published data ^22^, BMP4 levels were reduced in AM patients (Figure 3A) serving a good indicator for disease with an area under the curve (AUC) of 0.78 in the receiver operating characteristic (ROC) curve (Figure 3D). Both GREM1 and GREM2 concentrations were significantly elevated in the serum of AM patients when compared to healthy controls (Figure 3B-C), with GREM2 levels particularly increased in Cohort 2 (Supplementary data, Figure 3C). ROC curve analyses demonstrated that the dysregulation of the BMP4/GREM axis accurately distinguished AM patients from healthy controls (Figure 3D; Supplementary data, Table 3, Figure 3D) with the GREM2/BMP4 ratio (AUC 0.88, 95% CI 0.82 – 0.94) showing the strongest discriminatory power (Figure 3D). Multivariable analysis showed that GREM2 serum concentration was strongly associated with disease severity in AM patients, significantly correlating with both LVEF and NT-proBNP (Figure 3E-F). Moreover, changes in GREM2 were significantly associated with increased levels of fibroblast-derived factor HGF (Figure 3E and G) and the T cell-attracting chemokines CXL9 and CXCL10 (Figure 3E, Supplementary data, Figure 3E-F). In addition, HGF and GREM2 correlated strongly with the duration of hospitalisation in Cohort 1, serving as an additional indicator for clinical severity (Supplementary data, Figure 3G-I).

**Figure 3.**
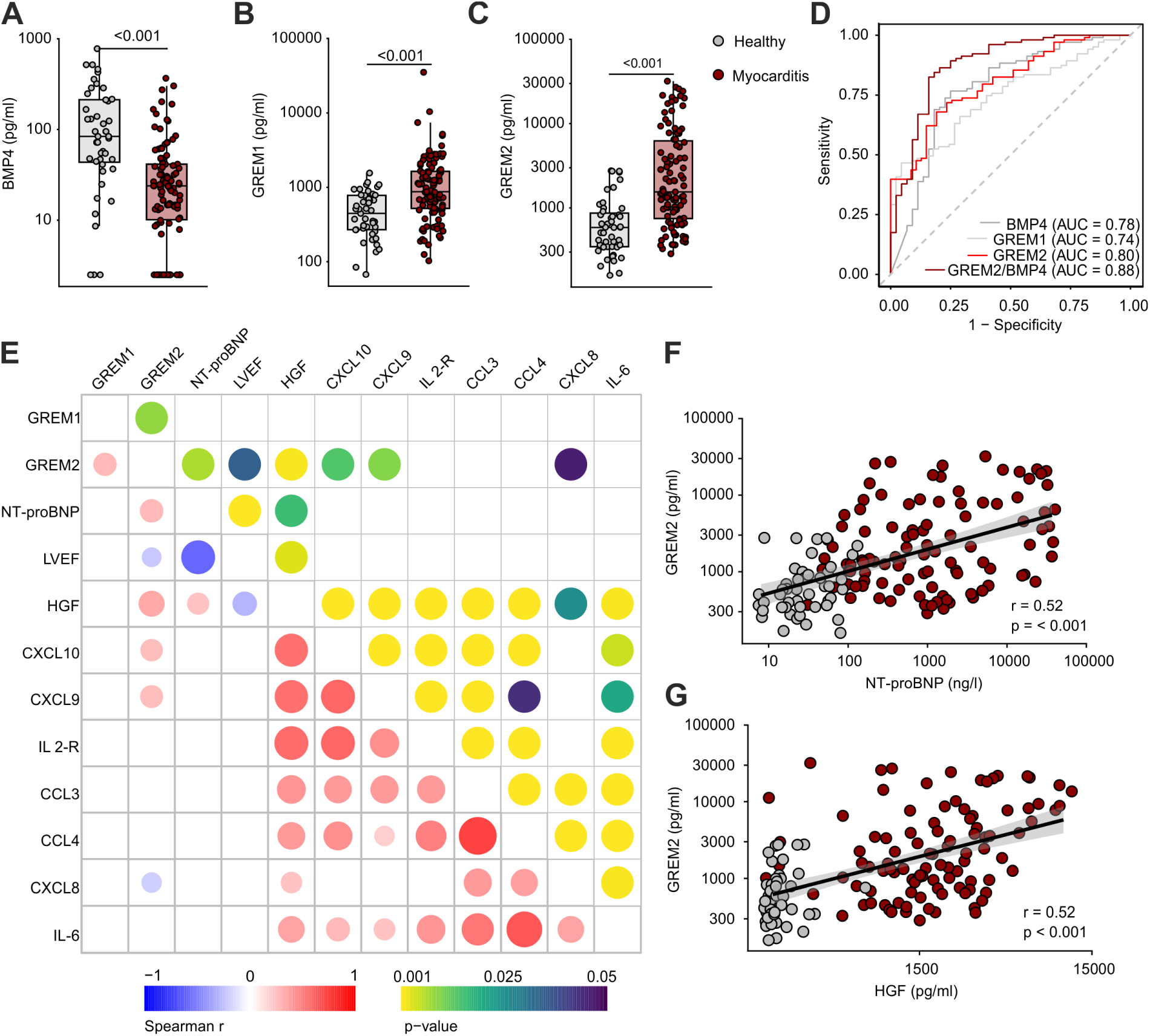
Association of BMP4/GREM2 dysregulation with disease severity in acute myocarditis. *(A-C)* Serum levels of BMP4 *(A)*, GREM1 *(B)* and GREM2 *(C)* in AM patients (n = 103) compared to healthy controls (n = 47). *(D)* ROC curve for BMP4, GREM1, GREM2 and the GREM2/BMP4 ratio to discriminate AM patients from healthy controls. *(E)* Multivariable correlation matrix showing the relationship between inflammatory cytokines, components of the BMP4-Gremlin axis and clinical parameters (NT-proBNP, LVEF) in AM patients *(F-G).* Correlation analyses of GREM2 levels and NT-proBNP *(F)* and HGF *(G)*. *(A-C, F-G)* Dots represent individual patients, *(A-C)* box and whiskers indicate minimum to maximum values, median, and interquartile range. (E) Bubble size in the lower triangle reflects the magnitude of correlation, bubble size in the upper triangle indicates the magnitude of significance. Bubble colour in the lower triangle denotes positive (red) and negative (blue) correlation, in the upper triangle, colour from blue to yellow represents increasing statistical significance. *(F-G)* Linear regression lines with 95% confidence interval (grey shading). Statistical analyses were performed using the Mann-Whitney U test for *(A-C)* and non-parametric Spearman correlation for *(E-G)*. P-values were corrected for multiple testing using the Benjamini-Hochberg method. AM, acute myocarditis, BMP4, bone morphogenic protein 4, GREM1, Gremlin-1, GREM2, Gremlin-2, ROC, Receiver operating characteristic, AUC, area under the curve, NT-proBNP, N-terminal pro-B-type natriuretic peptide, LVEF, left ventricular ejection fraction, HGF, hepatocyte-growth factor, IL-2R, interleukin-2 receptor

To assess whether the BMP4-GREM dysregulation is an exclusive feature of acute myocardial inflammatory disease, we performed a cross-disease comparison between patients with AM and arrhythmogenic cardiomyopathy (ACM) (Supplementary data, Table 6). Despite comparable heterogeneity in cardiac function, ACM patients exhibited only moderate elevation of NT-proBNP and Troponin I serum levels (Supplementary data, Figure 4A-D) with no substantial expression of inflammatory markers (Supplementary data, Figure 4E-G). The pattern of BMP4/GREM dysregulation in ACM patients was comparable to AM patients with reduction in BMP4 and increase in GREM1 concentration, while GREM2 levels were only moderately increased (Supplementary data, Figure 5H-J). In sum, these data underscore that dysregulation of the BMP4/GREM axis in AM is closely linked to T cell-associated immune activation and indicate that GREM2 functions as the dominant inhibitor of BMP4-mediated signalling during acute inflammatory processes in the myocardium.

### Fibroblast-T cell crosstalk drives immuno-clinical phenotype distinction

The finding that T cell-driven inflammatory signatures and fibroblast-associated remodelling processes showed significant association with disease severity suggested that the disease spectrum can be captured by a specific immuno-clinical profile. To this end, we used a set of machine-learning approaches including Uniform Manifold Approximation Projection (UMAP) as unsupervised dimensionality reduction technique to allow for unbiased clustering of key parameters identified from the immuno-proteomic and clinical data analyses (Supplementary data, Figure 5A-B). The algorithm identified two distinct immuno-clinical clusters (Figure 4A) with patients from Cohort 2 predominantly represented in cluster B (Figure 4B), reflecting the more severe clinical manifestations observed in this cohort (Table 1). Accordingly, cluster B was characterized by significantly impaired cardiac function and markedly elevated NT-proBNP levels (Supplementary data, Figure 5C–D), defining the clusters as mild versus severe immuno-clinical phenotypes. Indeed, patients with a severe phenotype showed significantly higher T-cell activation and fibroblast-associated signatures when compared to those with a mild phenotype (Supplementary data, Figure 5E-J). Patients with a severe immuno-clinical phenotype were significantly older (median age 39 vs. 28 years, p= 0.015), whereas sex distribution did not differ between phenotypes (Supplementary data, Table 5). Notably, a substantially higher percentage of patients with a severe immuno-clinical phenotype required immunosuppressive therapy (49% vs. 1.9%, p < 0.001) and mechanical circulatory support (22% vs. 0%, p < 0.001).

**Figure 4.**
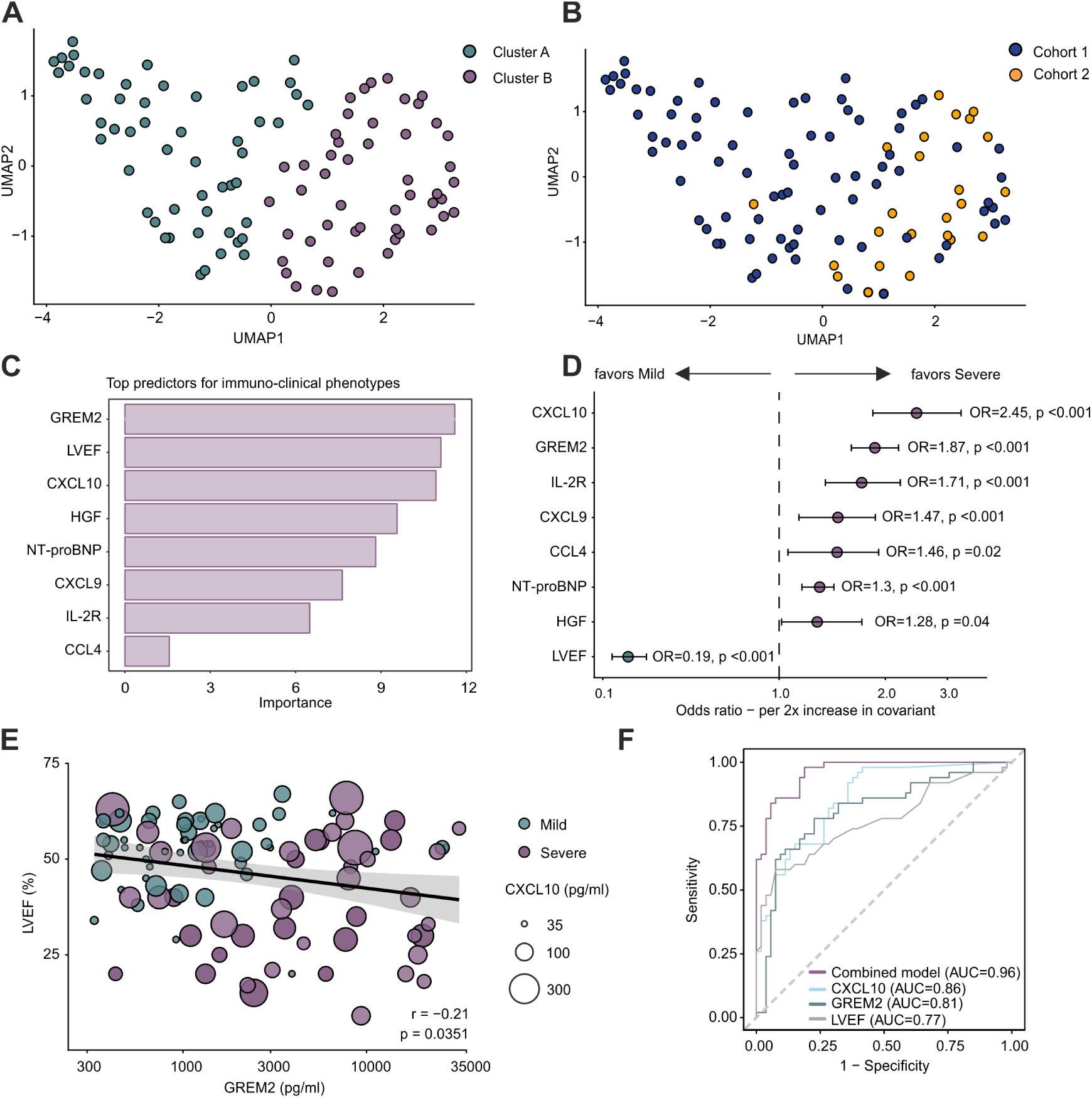
Identification of immuno-clinical phenotypes in acute myocarditis. *(A)* UMAP showing clustering of immuno-clinical phenotypes in patients with AM (n = 103). *(B)* UMAP illustrating the cohort distribution of AM patients across identified clusters (Cohort 1 n = 79, Cohort 2 n = 24) *(C)* Variable importance score of clinical and immunological parameters contributing to the discrimination of immuno-clinical phenotypes as determined by random forest analysis. *(D)* Forest plot displaying direction and magnitude of associations between preselected parameters with immuno-clinical phenotypes, expressed as OR for a 2-fold increase in the respective covariate in a multivariate ridge regression model. *(E)* Correlation analysis between GREM2 and LVEF, visually stratified by immuno-clinical phenotype. *(F)* ROC curve for CXCL10, GREM2, LVEF and a combined model incorporating CXCL10, GREM2 and LVEF parameters to distinguish between mild and severe immuno-clinical phenotype. *(A-B, E)* Dots represent individual patients. *(E)* Linear regression line with 95% confidence interval (grey shading), bubble size reflects CXCL10 levels, and bubble colour denotes immuno-clinical phenotypes and healthy controls. Statistical analyses were performed using unsupervised spiral clustering for *(A-B)*, random forest analysis was applied in *(C)*, multivariate ridge regression in *(D)* and non-parametric Spearman correlation for *(E).* P-values were derived from bootstrap resampling in *(D)*. UMAP, Uniform Manifold Approximation and Projection, GREM2, Gremlin-2, LVEF, left ventricular ejection fraction, HGF, hepatocyte-growth factor, NT-proBNP, N-terminal pro-B-type natriuretic peptide, IL-2R, interleukin-2 receptor, OR, odds ratio, AUC, area under the curve

To identify the key parameters driving the distinction of immuno-clinical phenotypes, we first applied the random forest algorithm, which identified GREM2, LVEF and CXCL10 as the most important predictors for phenotype specification (Figure 4C). Furthermore, ridge regression was used to quantify effect sizes and the direction of associations between individual parameters and phenotypes. The strongest covariance indicating a severe immuno-clinical phenotype were elevations in serum concentration of CXCL10 and GREM2 (CXCL10, OR 2.45, 95% CI: 1.86 – 3.31, p<0.001; GREM2, 1.83, 95% CI: 1.54 – 2.12, p<0.001). In contrast, preserved LVEF favoured a mild immuno-clinical phenotype (LVEF, OR 0.19, 95% CI: 0.11 – 0.29, p<0.001; Figure 4D; Supplementary data, Table 6). Correlation analysis highlighted the significant association of GREM2 and CXCL10 with clinical severity (Figure 4E; Supplementary data, Figure 5K). ROC curve analysis demonstrated that classification based on the key factors GREM2, CXCL10 and LVEF enabled highly accurate phenotype discrimination (Figure 4F; Supplementary Figure 5L). Notably, the combined use of GREM2, CXCL10 and LVEF confirmed the validity of the model and achieved excellent performance (AUC 0.96, 95% CI 0.93 – 0.99) in discriminating clinical severity in AM (Figure 4F; Supplementary data, Table 7), whereas the inclusion of additional parameters only marginally improved the AUC to 0.98 (95% CI 0.96 – 01.00) (Supplementary data, Figure 4L). Collectively, these findings underscore the diagnostic value of combining measurements of the T cell chemokine CXCL10 and the modulator of fibroblast activity GREM2 in conjunction with LVEF as discriminator for a severe immuno-clinical phenotype in AM.

## Discussion

The serological analysis of AM patients presented here shows that increased T cell-driven immune responses and dysregulated fibroblast activity are closely linked to clinical severity. The combination of the T cell chemoattractant CXCL10 and the modulator of fibroblast activity GREM2 together with the clinical standard parameter LVEF emerged as the strongest predictor of severe AM manifestation. These findings underpin the critical role of T cell-fibroblast interactions in the disease process and further highlight that T cell activation determines the degree of fibroblast-driven remodelling of the myocardium and concomitant impairment of cardiac function.

The core data of this study support the notion that T cell infiltration in the myocardium of AM patients is mediated by the chemokine receptor CXCR3, which binds the chemokines CXCL9 and CXCL10 ^35^. T cell accumulation in inflamed tissues relies to a large extent on the production of CXCL9 and CXCL10 by activated macrophages and fibroblasts ^22,36,37^. It is thus likely that the increase of CXCL9 and CXCL10 in the serum of AM patients is a consequence of macrophage and fibroblast activation, which is a hallmark of myocardial inflammation ^18,22,29,38^. Indeed, antibody-mediated blocking of CXCL10 or CXCR3 has been shown to ameliorate myocardial immune cell infiltration in mouse models of myocarditis ^39,40^. The association of increased CXCL10 serum levels in patients with severe heart failure symptoms underscores that T cell-attracting chemokines serve as indicators of inflammatory myocardial disease activity ^41,42^. Future studies should therefore assess the value of an extended set of serum biomarkers reflecting T cell activity, such as CXCL9 and IL-2R, to identify episodes of myocardial inflammation in cardiomyopathy and heart failure patients.

The dysregulated crosstalk between activated T cells and fibroblasts in the inflamed myocardium is one of the major pathways leading to cardiomyocyte damage ^19,21,22^. In particular, the presence of activated T cells in the myocardium precipitates profound phenotypical changes of cardiac fibroblasts with the acquisition of an inflammatory state ^18,22,25,43^ with IFN-γ and IL-17 serving as key drivers of this process^6,9,10,21^. The accumulation of activated T cells and inflammatory macrophages in the myocardium cause a reduction in homeostatic BMP receptor signalling ^22^. The association between dysregulation of the BMP4-GREM axis and T cell-driven myocardial inflammation in AM patients, as demonstrated in this study, underscores the critical role of BMP4-mediated signalling in fibroblasts for maintaining tissue homeostasis. The increased GREM1 serum concentration in AM and ACM patients supports the notion that GREM1 is a general marker for fibroblast activation associated with inflammatory and fibrotic processes across multiple tissues ^44–46^. In contrast, GREM2 emerged as a BMP4 antagonist more closely linked to T cell-mediated cardiac dysfunction. As inflammatory processes and the restoration of tissue homeostasis depend on multi-level regulatory circuits ^47,48^, we consider it likely that the control of BMP receptor signalling by GREM1 and GREM2 serves as a tuneable system to govern both inflammatory and fibrotic processes in the myocardium.

The current disease classification of AM patients relies largely on clinical parameters, which lack specificity for myocardial inflammation and provide limited insight into the underlying immunopathology ^3^. For example, serum or plasma levels of Troponin I do not correlate with the degree of cardiac dysfunction observed in AM patients, while subtle changes in the neutrophil-to-lymphocyte ratio have been associated with disease severity and adverse outcome ^49,50^. Moreover, these studies underscore that CRP provides limited value as indicator for disease severity in AM ^49,50^, a finding confirmed in the present study. In addition, the data presented here demonstrate that targeted immuno-profiling in combination with the functional parameters LVEF and NT-proBNP, is most useful to stratifying disease severity in AM. The improved diagnostic accuracy and mechanistic insights into key processes involved in disease mechanisms may help to broaden the currently limited therapeutic options for AM. Indeed, despite the lack of robust evidence, glucocorticoids remain the main first-line immunosuppressive therapy for severe (fulminant) myocarditis ^3^. The increasing availability of targeted immuno-modulating therapies requires a detailed understanding of cardiopathogenic immune responses to enable tailored immuno-therapeutical treatment in AM ^2^. The distinct inflammatory signatures identified in this study may guide patient stratification for targeted immunotherapy in future clinical trials. Moreover, this panel of inflammatory parameters could be used for longitudinal monitoring of the disease and may help identify patients at risks for a chronic or recurrent disease course.

## Study limitations

Our study identified CXCL10 and GREM2 as novel biomarkers for clinical severity in AM, underscoring the relevance of T cell-fibroblast interactions as a key component of myocardial inflammation. However, as the present study is based on circulating biomarkers, future work is warranted to directly validate these interactions within the inflamed myocardium. In addition, a subset of patients received immunosuppressive therapy prior to study enrolment. Thus, a potential effect of immunomodulatory treatment on the serological immune profiles cannot be fully excluded. Nevertheless, the preservation of inflammatory cytokine and chemokine signatures across both cohorts (i.e., 13% in Cohort 1 vs. 63% Cohort 2 received immunosuppressive treatment), indicates that the identified immuno-clinical phenotypes are robust and reliable.

## Conclusions and Future Perspectives

This study highlights a systemic T cell activation signature as diagnostic hallmark of AM. In addition, dysregulation of fibroblast-derived tissue cytokines serves as an indicator for distinct immuno-clinical phenotypes in myocardial inflammatory disease. These findings highlight the potential of serological immune profiling to capture inflammatory mechanisms driving myocardial injury and to improve the identification of patients at risk of severe AM.

The immuno-clinical phenotypes identified in our study provide a diagnostic framework for stratifying AM patients by integrating clinical manifestation with distinct inflammatory signatures. This approach provides a strong foundation for precision medicine strategies aimed to guide the selection of immunomodulatory therapies and enabling the monitoring of treatment responses. Moreover, the identification of the fibroblast-derived factor GREM2 as an indicator of disease severity highlights a potential therapeutic target to ameliorate inflammation-induced cardiac remodelling rather than broadly suppressing the systemic immune response. Future longitudinal follow-up studies of these immuno-clinical phenotypes may provide important insights into their prognostic value for disease progression and clinical outcome.

## Data Availability

The data supporting the findings of this study are available from the corresponding author upon reasonable request.

## Acknowledgments

The authors thank the study coordination team at the Zürich University Heart Center and at the Cardiology Department of the Cantonal Hospital St. Gallen for their valuable support in the ImmpathCarditis study. We further express our gratitude to Dr. rer. nat. Patrick Münzer for his assistance in coordinating sample handling for Cohort 2. We also gratefully acknowledge all patients who participated in the included studies, as well as all individuals enrolled in the healthy cohort.

## Abbreviation and Acronyms

ACM: Arrhythmogenic cardiomyopathy
AM: Acute Myocarditis
BMP4: Bone morphogenic protein 4
CMR: Cardiac magnetic resonance
CXCL: C-X-C motif chemokine ligand
EMB: Endomyocardial biopsy
GREM1: Gremlin-1
GREM2: Gremlin-2
HGF: Hepatocyte-growth factor
ICI: Immune-checkpoint inhibitor
IL: Interleukin
IL-2R: Interleukin-2 receptor
LVEF: Left ventricular ejection fraction

