## Supplementary Figures and Tables for "T-cell activation and fibroblastic BMP4-Gremlin dysregulation indicate disease severity in acute myocarditis"

Supplementary Material

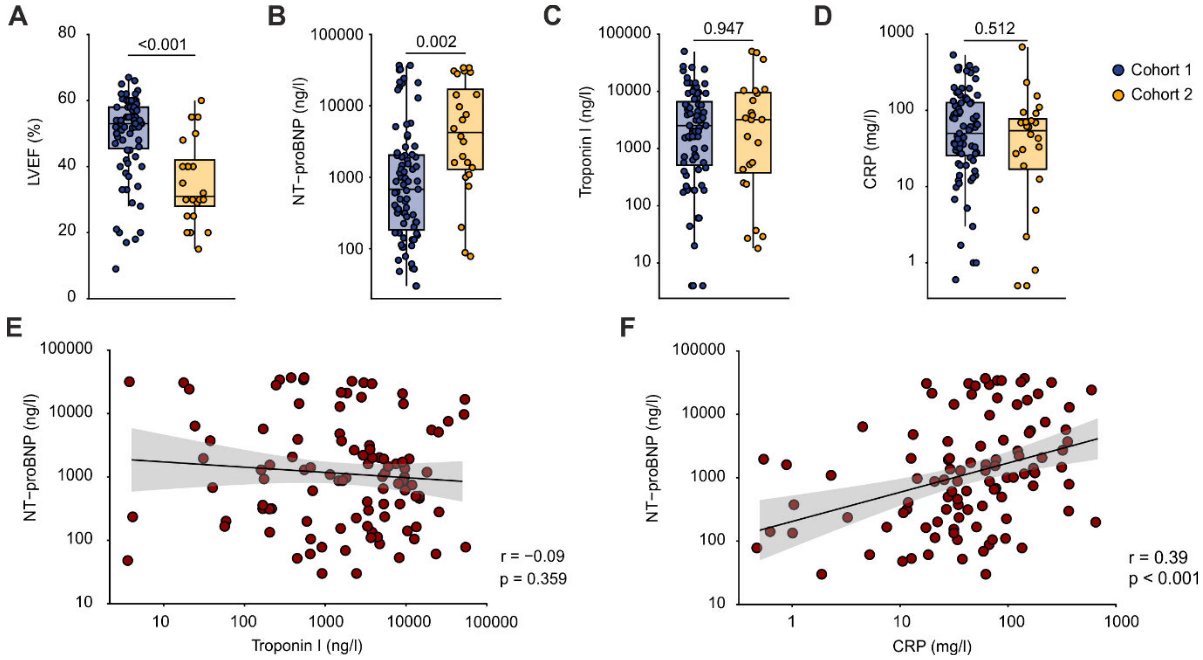


**Supplementary Figure 1: Clinical parameters in AM cohorts.** *(A)* LVEF compared between Cohort 1 and Cohort 2. *(B-D)* Comparison of peak values of routine blood parameters during hospitalization in Cohort 1 vs. Cohort 2: *(B)* NT-proBNP, *(C)* Troponin I, *(D)* CRP. *(E-F)* Correlations between NT-proBNP and other routine blood parameters: *(E)* Troponin I *(F)* and CRP. *(A-F)* Dots represent individual patients; *(A-D)* box and whiskers indicate minimum to maximum values, median, and interquartile range, *(E-F)* linear regression lines with 95% confidence interval (grey shading). Statistical analyses were performed using the Mann-Whitney U-test for *(A-D)* and non-parametric non-parametric Spearman correlation for *(E-F)*. P-values were corrected for multiple testing using the Benjamini-Hochberg method. AM, acute myocarditis, LVEF, left ventricular ejection fraction, NT-proBNP, N-terminal pro-B-type natriuretic peptide, CRP, C-reactive protein.

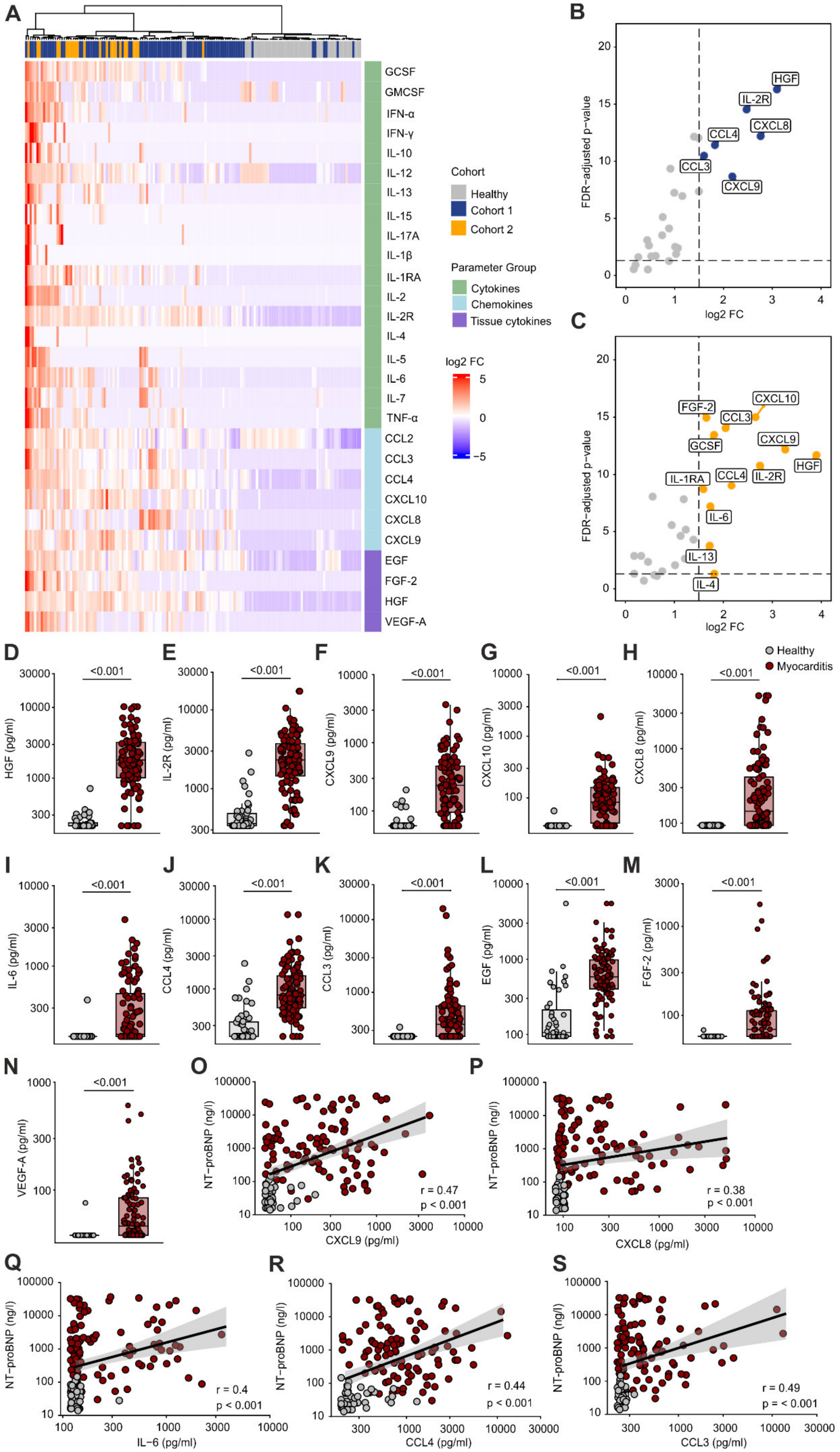

A
B
C
D
E
F
G
H
I
J
K
L
M
N
O
P
Q
R
S
GCSF
GMCSF
IFN-α
IFN-γ
IL-10
IL-12
IL-13
IL-15
IL-17A
IL-1β
IL-1RA
IL-2
IL-2R
IL-4
IL-5
IL-6
IL-7
TNF-α
CCL2
CCL3
CCL4
CXCL10
CXCL8
CXCL9
EGF
FGF-2
HGF
VEGF-A
Cohort
Healthy
Cohort 1
Cohort 2
Parameter Group
Cytokines
Chemokines
Tissue cytokines
log2 FC
FDR-adjusted p-value
Healthy
Myocarditis
<0.001
HGF (pg/ml)
IL-2R (pg/ml)
CXCL9 (pg/ml)
CXCL10 (pg/ml)
CXCL8 (pg/ml)
IL-6 (pg/ml)
CCL4 (pg/ml)
CCL3 (pg/ml)
EGF (pg/ml)
FGF-2 (pg/ml)
VEGF-A (pg/ml)
NT-proBNP (ng/l)
r = 0.47
p < 0.001
CXCL9 (pg/ml)
r = 0.38
p < 0.001
CXCL8 (pg/ml)
r = 0.4
p < 0.001
IL-6 (pg/ml)
r = 0.44
p < 0.001
CCL4 (pg/ml)
r = 0.49
p = < 0.001
CCL3 (pg/ml)

**Supplementary Figure 2: Conserved inflammatory signatures across two independent AM cohorts.** *(A)* Serum levels of 28 cytokines, chemokines and tissue-derived cytokines measured by Luminex in AM patients (Cohort 1 n = 79, Cohort 2 n = 24) and healthy controls (n = 47). *(B-C)* Volcano plot with differentially expressed proteins in *(B)* Cohort 1 and *(C)* Cohort 2 compared with healthy controls. *(D-K)* Differentially expressed inflammatory molecules in AM patients compared with healthy controls: *(D)* HGF, *(E)* IL-2R, *(F)* CXCL9, *(G)* CXCL10, *(H)* CXCL8, *(I)* IL-6, *(J)* CCL4 and *(K)* CCL3. *(L-N)* Expression of fibroblasts-derived tissue cytokines in AM patients compared with healthy controls: *(L)* EGF, *(M)* FGF-2, *(N)* VEGF-A. *(O-S)* Correlation analyses between NT-proBNP and inflammatory molecules in AM patients and healthy controls: *(O)* CXCL9, *(P)* CXCL8, *(Q)* IL-6, *(R)* CCL4, *(S)* CCL3. *(A)* High protein expression is shown in red and low expression in blue, indicated as log2 FC. *(B-C)* Coloured dots indicate inflammatory molecules expressed >1.5 log2 FC and p <0.05 in AM patients *(D-N)* Box and whiskers indicate minimum to maximum values, median, and interquartile range. *(D-S)* Dots represent individual patients. *(O-S)* Linear regression lines with 95% confidence interval are shown (grey shading). Statistical analyses were performed using Ward´s unsupervised hierarchical clustering method for *(A)*, Mann-Whitney U-test for *(B-N)* and non-parametric Spearman correlation for *(O-S)*. P values were corrected for multiple testing using the Benjamini-Hochberg method. AM, acute myocarditis, FC, fold change, G-CSF, granulocyte colony-stimulating factor, GM-CSF, granulocyte-macrophage colony-stimulating factor, IFN-α, interferon alpha, IFN-γ, interferon gamma, IL-1β, interleukin-1 beta, IL-1RA, interleukin-1 receptor antagonist, IL-2R, interleukin-2 receptor, TNF-α, tumor necrosis factor alpha, EGF, epidermal growth factor, HGF, hepatocyte-growth factor, VEGF-A, vascular endothelial growth factor A, FDR, false discovery rate, NT-proBNP, N-terminal pro-B-type natriuretic peptide

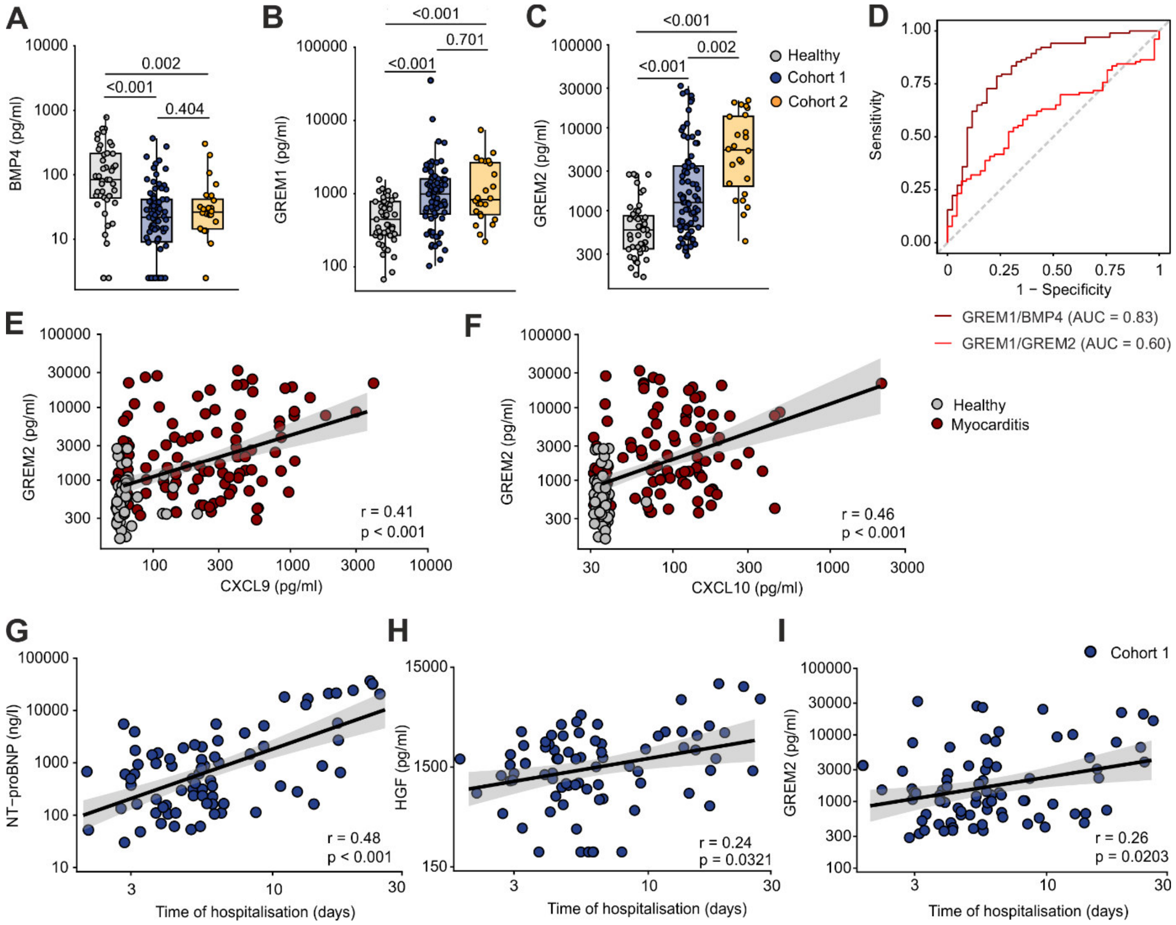


**Supplementary Figure 3: Dysregulation of the fibroblastic BMP4/GREM2 axis in acute myocarditis.** (*A-C)* Serum levels of BMP4 *(A)*, GREM1 *(B)* and GREM2 *(C)* in two independent AM cohorts (Cohort 1 n = 79, Cohort 2 n = 24) and healthy controls (n = 47). *(D)* ROC curve for GREM1/BMP4 ratio and GREM1/GREM2 ratio. *(E-F)* Correlation analyses between GREM2 and *(E)* CXCL9 and *(F)* CXCL10. *(G-I)* Correlation analyses between hospitalisation time and *(G)* NT-proBNP, *(H)* HGF, and *(I)* GREM2 in Cohort 1. *(A-C, E-I)* Dots represent individual patients. *(A-C)* Box and whiskers indicate minimum to maximum values, median, and interquartile range. *(E-I)* Linear regression lines with 95% confidence interval (grey shading). Statistical analyses were performed using the Mann-Whitney U test for *(A-C)* and non-parametric Spearman correlation for *(E-I)*. P-values are corrected for multiple testing using the Benjamini Hochberg correction. AM, acute myocarditis, BMP4, bone morphogenic protein 4, GREM1, Gremlin-1, GREM2, Gremlin-2, ROC, Receiver operating characteristic, AUC, area under the curve, NT-proBNP, N-terminal pro-B-type natriuretic peptide, HGF, hepatocyte-growth factor

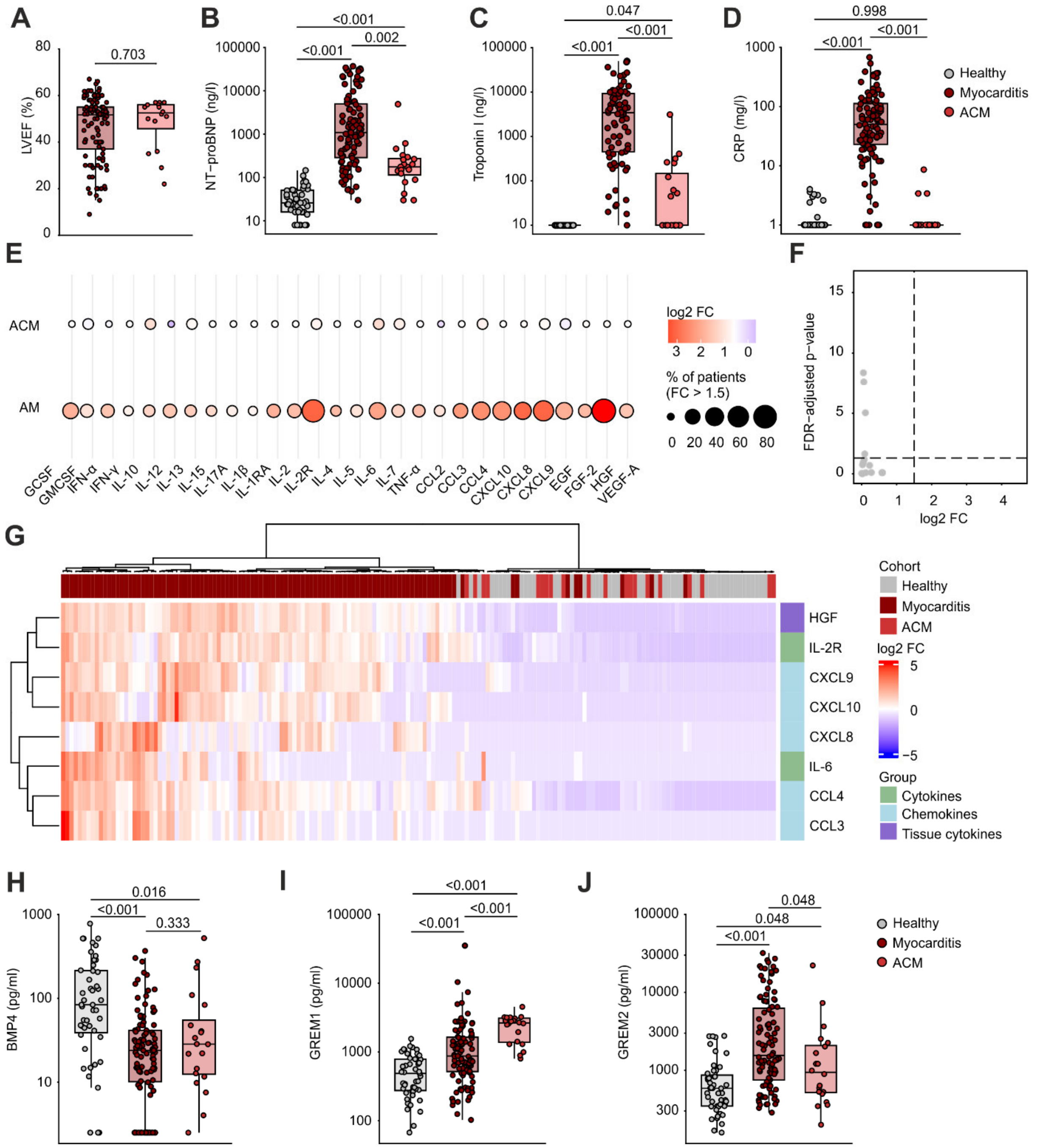


**Supplementary Figure 4: BMP4 and GREM1/2 dysregulation in arrhythmogenic cardiomyopathy (ACM).** *(A)* LVEF in ACM patients (n = 20) compared to AM patients (n = 103). *(B-D)* Routine blood parameters in ACM patients, AM patients (peak values) and healthy controls (n = 47): *(B)* NT-proBNP, *(C)* Troponin I, *(D)* CRP. *(E)* Bubble plot showing expression pattern of cytokines, chemokines and tissue-derived cytokines in ACM patients and AM patients. *(F)* Volcano plot with differentially expressed proteins in ACM patients compared to healthy controls. *(G)* Heatmap of inflammatory serum molecules in ACM patients, AM patients and healthy controls. *(H-J)* Serum levels of BMP4 *(H)*, GREM1 *(I)* and GREM2 *(J)* in ACM patients compared to AM patients and healthy controls. *(A-D, H-J)* Dots represent individual patients, box and whiskers indicate minimum to maximum values, median, and interquartile range. *(E)* Bubble size represents the abundance of inflammatory molecules >1.5 log2 FC compared to healthy controls and bubble colour indicates magnitude of log2 FC. *(F)* Grey dots represent non-significant elevated inflammatory molecules with < 1.5 log2 FC. *(G)* High protein expression is shown in red and low expression in blue, indicated as log2 FC. Statistical analyses were performed using the

Mann-Whitney U-test for *(A, E-F),* the Kruskal-Wallis test followed by Dunn`s post-hoc test for *(B-D, H-J)* and Ward´s unsupervised hierarchical clustering method for *(G).* P-values were corrected for multiple testing using the Benjamini-Hochberg method. ACM, arrhythmogenic cardiomyopathy, AM, acute myocarditis, LVEF, left ventricular ejection fraction, NT-proBNP, N-terminal pro-B-type natriuretic peptide, CRP, C-reactive protein, FC, fold change, G-CSF, granulocyte colony-stimulating factor, GM-CSF, granulocyte-macrophage colony-stimulating factor, IFN-α, interferon alpha, IFN-γ, interferon gamma, IL-1β, interleukin-1 beta, IL-1RA, IL-1RA, interleukin-1 receptor antagonist, IL-2R, interleukin-2 receptor, TNF-α, tumor necrosis factor alpha, EGF, epidermal growth factor, HGF, hepatocyte-growth factor, VEGF-A, vascular endothelial growth factor A, FDR, false discovery rate, BMP4, bone morphogenic protein 4, GREM1, Gremlin-1, GREM2, Gremlin-2

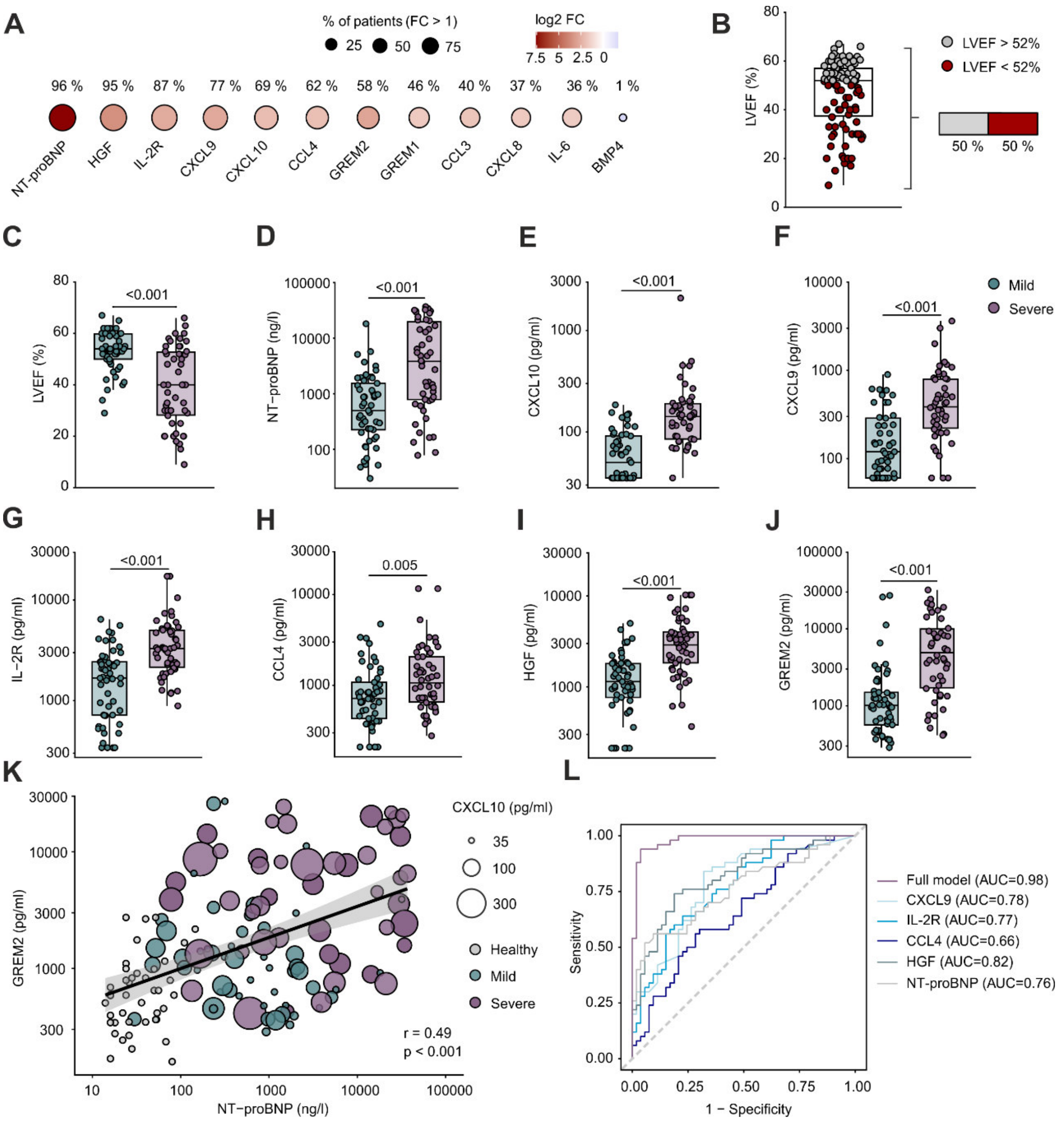


**Supplementary Figure 5: Inflammatory and clinical parameters across immuno-clinical phenotypes.** *(A)* Bubble plot of expression patterns and relative abundances of differentially expressed inflammatory molecules and clinical parameters in AM patients (n= 103) compared with healthy controls (n= 47). Parameters with log2 FC >1 in more than 50% of AM patients were used for subsequent cluster analysis. *(B)* Prevalence of reduced LVEF (<52%) in AM patients. LVEF was included as a clinically relevant parameter for subsequent cluster analysis *(C-J)* Expression profile of inflammatory molecules and clinical parameters in the two identified immuno-clinical phenotypes (Mild n=53, Severe n= 50): *(C)* LVEF, *(D)* NT-proBNP, *(E)* CXCL10, *(F)* CXCL9, *(G)* IL-2R, *(H)* CCL4, *(I)* HGF and *(J)* GREM2. *(K)* Correlation analysis visually stratified by immuno-clinical phenotype between GREM2 and NT-proBNP. *(L)* ROC curve for CXCL9, IL-2R, CCL4, HGF, NT-proBNP and for a full model incorporating all eight parameters used in the unsupervised clustering analysis (CXCL9, IL-2R, CCL4, HGF, NT-proBNP, CXCL10, GREM2, LVEF) to discriminate between immuno-clinical phenotypes. *(B-K)* Dots represent individual patients. *(B-J)* Box and whiskers indicate minimum to maximum values, median, and interquartile range. *(K)* Linear regression line with 95% confidence interval (grey shading), bubble size corresponds to CXCL10 levels, and bubble colours denote immuno-clinical phenotypes and healthy controls. Statistical analyses were performed using the Mann-Whitney U-test with for *(A, C-J)* and non-parametric Spearman correlation for *(K)*. P-values are corrected for multiple testing using the Benjamini Hochberg correction. AM, acute myocarditis, NT-proBNP, N-terminal pro-B-type natriuretic peptide, HGF, hepatocyte-growth factor, IL-2R, interleukin-2 receptor, GREM2, Gremlin-2, GREM1, Gremlin-1, BMP4, bone morphogenic protein 4, LVEF, left ventricular ejection fraction, FC, fold change, ROC, Receiver operating characteristic, AUC, area under the curve

**Supplementary Table 1:** Detection limits of inflammatory molecules measured by Luminex.

| Analyte | LLOQ (pg/ml) | ULOQ (pg/ml) |
|---|---|---|
| FGF-2 | 58.36 | 3155.00 |
| IL-1β | 137.89 | 6250.00 |
| G-CSF | 940.16 | 45400.00 |
| IL-10 | 305.31 | 7180.00 |
| IL-13 | 196.48 | 9450.00 |
| IL-6 | 133.36 | 6220.00 |
| IL-12 | 96.56 | 5280.00 |
| CCL11 | 67.58 | 3905.00 |
| IL-17A | 368.28 | 15740.00 |
| CCL3 | 249.30 | 14115.00 |
| GM-CSF | 85.47 | 4140.00 |
| CCL4 | 209.53 | 11630.00 |
| CCL2 | 289.84 | 15515.00 |
| IL-15 | 247.89 | 10190.00 |
| EGF | 94.38 | 5465.00 |
| IL-5 | 174.22 | 5570.00 |
| HGF | 211.72 | 10255.00 |
| VEGF-A | 38.05 | 2275.00 |
| IFN-γ | 46.88 | 2319.00 |
| IFN-α | 133.59 | 8225.00 |
| IL-1RA | 1542.66 | 95115.00 |
| TNF-α | 52.50 | 2560.00 |
| IL-2 | 152.73 | 8655.00 |
| IL-7 | 217.89 | 10495.00 |
| CXCL10 | 35.23 | 2085.00 |
| IL-2R | 340.78 | 17210.00 |
| CXCL9 | 60.39 | 3650.00 |
| IL-4 | 562.42 | 32940.00 |
| CXCL8 (IL-8) | 93.44 | 5135.00 |

**Supplementary Table 1:** Detection limits of inflammatory molecules measured by Luminex. LLOQ: lower limit of quantification, ULOQ: upper limit of quantification, IL: Interleukin, CCL: CC chemokine ligand, CXCL: CXC chemokine ligand, FGF-2: fibroblast growth factor, IL-1β, interleukin-1 beta, G-CSF: granulocyte colony-stimulating factor, GM-CSF: granulocyte-macrophage colony-stimulating factor, EGF: epidermal growth factor, HGF: hepatocyte growth factor, VEGF-A: vascular endothelial growth factor A, IFN-γ, interferon gamma, IFN-α, interferon alpha, IL-1RA: interleukin-1 receptor antagonist, TNF-α, tumor necrosis factor alpha, IL-2R: interleukin-2 receptor

**Supplementary Table 2:** Demographics of healthy controls and acute myocarditis patients.

| Characteristic | Healthy (n = 47) | Acute Myocarditis (n = 103) | p-value |
|---|---|---|---|
| Sex | | | 0.2 |
| Female (%) | 14 (30%) | 21 (20%) | |
| Male (%) | 33 (70%) | 82 (80%) | |
| Age | 30 (22 - 61) | 34 (17 - 73) | 0.7 |
| **Recruitment Centre[a]** | | | |
| ZH | 0 (0%) | 64 (62%) | |
| SG | 47 (100%) | 15 (15%) | |
| TU | 0 (0%) | 24 (23%) | |

**Supplementary Table 2:** Demographic characteristics of healthy controls and acute myocarditis patients. Healthy controls were sex- and age-matched. Continuous variables are presented as median (min-max), and categorical variables as counts. Statistical comparisons were performed using the Mann-Whitney U-test for continuous variables and Pearson's Chi-squared test or Fisher's exact test for categorical data. [a]Recruitment centres: ZH: Department of Cardiology, University Hospital Zurich, Zurich, Switzerland, SG: Department of Cardiology, Cantonal Hospital St. Gallen, St. Gallen, Switzerland, TU: Department of Cardiology and Angiology, Eberhard Karls University of Tübingen, Tübingen, Germany

**Supplementary Table 3:** Performance of the BMP4-GREM dysregulation to distinguishing AM patients from healthy controls.

| **Parameter** | **AUC** | **95% CI** |
|---|---|---|
| BMP4 | 0.78 | 0.69 – 0.87 |
| GREM1 | 0.74 | 0.66 – 0.82 |
| GREM2 | 0.80 | 0.73 – 0.87 |
| GREM2/BMP4 | 0.88 | 0.81 – 0.94 |
| GREM1/BMP4 | 0.83 | 0.76 – 0.91 |
| GREM1/GREM2 | 0.60 | 0.51 – 0.70 |

**Supplementary Table 3:** Receiver operating characteristic (ROC) curves of BMP4, Gremlin-1 and Gremlin-2 showing accuracy to distinguish between AM patients from healthy controls. BMP4, bone morphogenic protein 4, AM, acute myocarditis, AUC, area under the curve, CI, confidence interval

**Supplementary Table 4:** Characteristics of the arrhythmogenic cardiomyopathy (ACM) Cohort

| **Characteristics** | **ACM (**n= 20) |
|---|---|
| Sex | |
| Female (%) | 12 (60%) |
| Male (%) | 8 (40%) |
| Age | 51 (18 - 72) |
| ICD | 13 (65%) |
| **Cardiac function** | |
| LVEF (%)* | 53 (22 - 57) |
| RV fac (%)* | 24 (15 - 39) |
| LVEDVi (ml/$m^2$)* | 58 (39 - 96) |
| LV regional wall motion abnormalities* | 7 (35%) |
| **Mutations** | |
| DSG-2 | 3 (15%) |
| DSP | 6 (30%) |
| PKP-2 | 11 (55%) |
| **Comorbidities** | |
| Dyslipidaemia | 2 (10%) |
| Diabetes | 0 (0%) |
| Arterial Hypertension | 1 (5%) |

**Supplementary Table 4:** Patient characteristics of the ACM Cohort. Continuous variables are presented as median (min-max), and categorical variables as counts (%). ICD: implantable cardioverter-defibrillator, LVEF : Left ventricular ejection fraction, RV fac: Right ventricular fractional area change, LVEDVi: Left ventricular end-diastolic volume index, LV regional wall motion abnormalities: Left ventricular wall motion abnormalities, DSG-2: Desmoglein-2, DSP: Desmoplakin, PKP-2: Plakophillin-2. *Derived from the most recent echocardiographic assessment.

**Supplementary Table 5:** Characteristics of patients with mild and severe immuno-clinical phenotype.

| Characteristic | **Mild** (n= 53) | **Severe** (n= 50) | p-value |
|---|---|---|---|
| Age | 28 (17 - 73) | 39 (18 - 70) | 0.015 |
| Sex, Male (%) | 45 (85%) | 37 (74%) | 0.2 |
| **Recruitment Centre[a]** | | | |
| ZH | 41 (77%) | 23 (46%) | |
| SG | 11 (21%) | 4 (8.0%) | |
| TU | 1 (1.9%) | 23 (46%) | |
| **Clinical presentation** | | | |
| Dyspnoea | 9 (17%) | 20 (40%) | 0.015 |
| **Routine blood parameters** | | | |
| CRP (mg/dl) | 37 (1 - 388) | 61 (1 - 679) | 0.3 |
| Troponin I (ng/ml) | 3,302 (4 - 27,327) | 3,626 (18 - 50,000) | >0.9 |
| NT-proBNP (ng/ml) | 552 (30 - 18,150) | 3,735 (78 - 36,847) | <0.001 |
| **In-hospital management** | | | |
| Immunosuppression | 1 (1.9%) | 24 (49%) | <0.001 |
| Mechanical circulatory support | 0 (0%) | 11 (22%) | <0.001 |
| **Immunological parameters** | | | |
| CXCL10 (pg/ml) | 50 (35 - 185) | 142 (35 - 2,085) | <0.001 |
| CXCL9 (pg/ml) | 120 (60 - 896) | 386 (60 - 3,619) | <0.001 |
| HGF (pg/ml) | 1,143 (212 - 4,995) | 2,894 (367 - 10,255) | <0.001 |
| CCL4 (pg/ml) | 718 (210 - 4,733) | 1,067 (278 - 11,630) | 0.005 |
| IL-2R (pg/ml) | 1,677 (341 - 6,425) | 3,285 (886 - 17,210) | <0.001 |
| Gremlin-2 (pg/ml) | 1,010 (286 - 27,112) | 4,911 (413 - 31,976) | <0.001 |

**Supplementary Table 5:** Patient characteristics comparing mild and severe immuno-clinical phenotypes. Continuous variables are presented as median (min-max), and categorical variables as counts (%). Statistical comparisons were performed using the Mann-Whitney U-test for continuous variables and Pearson's Chi-squared test or Fisher's exact test for categorical data. HGF: Hepatocyte growth factor, IL-2R: Interleukin-2 receptor. [a]Recruitment centres: ZH: Department of Cardiology, University Hospital Zurich, Zurich, Switzerland, SG: Department of Cardiology, Cantonal Hospital St. Gallen, St. Gallen, Switzerland, TU: Department of Cardiology and Angiology, University of Tübingen, Tübingen, Germany

**Supplementary Table 6:** Ridge regression model for the identification of distinctive parameters in AM immuno-clinical phenotypes.

| Parameter | OR | 95 % CI | p-value |
|---|---|---|---|
| CXCL10 | 2.45 | 1.84 – 3.28 | < 0.001 |
| Gremlin-2 | 1.87 | 1.60 – 2.18 | < 0.001 |
| IL-2R | 1.71 | 1.35 – 2.21 | < 0.001 |
| CXCL9 | 1.47 | 1.14 – 1.87 | < 0.001 |
| CCL4 | 1.46 | 1.06 – 1.92 | < 0.001 |
| NT-proBNP | 1.30 | 1.16 – 1.43 | < 0.001 |
| HGF | 1.28 | 1.02 – 1.72 | 0.04 |
| LVEF | 0.19 | 0.13 – 0.31 | < 0.001 |

**Supplementary Table 6:** Statistical results of ridge regression model identifying most relevant parameters in immuno-clinical phenotypes in AM patients. Values apply to 2-fold increase of covariant. 95 % CI and p-values were estimated using 100 bootstrap resampling recapitulations. OR: odds ratio, CI: confidence interval, IL-2R: Interleukin-2 receptor, HGF: hepatocyte growth factor

**Supplementary Table 7:** Performance of immunological and clinical parameters distinguishing immuno-clinical phenotypes in AM patients.

| Parameter | AUC | 95% CI |
|---|---|---|
| CXCL10 | 0.86 | 0.79 – 0.93 |
| Gremlin-2 | 0.81 | 0.73 – 0.90 |
| LVEF | 0.77 | 0.68 – 0.86 |
| Combined model* | 0.96 | 0.93 – 0.99 |
| IL-2R | 0.77 | 0.68 – 0.86 |
| CXCL9 | 0.78 | 0.69 – 0.87 |
| CCL4 | 0.66 | 0.55 – 0.76 |
| NT-proBNP | 0.75 | 0.66 – 0.85 |
| HGF | 0.82 | 0.73 – 0.90 |
| Full model⁺ | 0.98 | 0.96 – 1.00 |

**Supplementary Table 7:** Receiver operating characteristic (ROC) curves of immunological and clinical parameters showing accuracy to distinguish between immuno-clinical phenotypes in AM patients. AM, acute myocarditis, AUC, area under the curve, CI, confidence interval, IL-2R: Interleukin-2 receptor, HGF: hepatocyte growth factor, *Combined model includes CXCL10, Gremlin-2 and LVEF, ⁺Full model includes all preselected parameters used for unsupervised clustering: CXCL10, Gremlin-2, LVEF, IL-2R, CXCL9, CCL4, NT-proBNP, HGF
